# The First Month of COVID-19 in Madagascar

**DOI:** 10.1101/2020.04.23.20076463

**Authors:** Stephan Narison

## Abstract

Using the officially published data and aware of the unclear source and insufficient number of samples, we present a first and (for the moment) unique attempt to study the spread of the pandemic COVID-19 in Madagascar. The approach has been tested by predicting the number of contaminated persons until 20 days after fitting the inputs data collected within 15 days using standard least, *χ*^2^-fit method. Encouraged by this first test, we add the new data collected within 30 days and give prevision until 33 days. We complete the analysis by using the updated data until 46 days. The data below 30 days show an approximate linear or quadratic polynomial increase of about (4-5) infected persons per day. A comparison with some other SI-like models is done. However, newer additional data collected until 46 days favours a cubic polynomial behaviour which signals an eventual near future stronger growth as confirmed by the data on the 48th day received after our study and our estimate including this new data for the next days. The analysis of the numbers of cured persons for 38 and 46 days shows behaviours similar to the ones of the infected persons. They indicate about 3 cured persons per day. These results for infected and cured persons may also be interpreted as the lowest values of the real cases due to the insufficient number of samples (about 3968 for 27 million inhabitants on 07/05/2020). Some social, economical and political impacts of COVID-19 and confinement for Madagascar and Worldwide are shortly discussed.

## 1. Introduction

• COVID-19 is scientifically named SRAS (Syndrome Respiraroire Aigu Severe) or SARS (Severe Acute Respiratory Syndrome)-CoV-2 or COVID-2-19 or COVID-19 in reference to the SRAS-CoV or Coronavirus pandemic (2002-2003) starting in China and propagated in 30 countries which has affected 8 000 persons and caused 774 deceased [1]. There is a slight difference between the two COVID in the genome structure of the virus thus the symbol 2.

• Another pandemic named MERS-CoV (2012-2013) or SRAS of the MIDDLE EAST has been found for the first time in Saudi Arabia and has affected 1 589 persons and caused 567 deceased in 26 countries.

• The CORONAVIRUS are of animal origins. An asymptomatic animal transmits to another ones and then to human. It was bats for SRAS-COV and MERS-COV which transmit the virus to masked palm civets for China and to camels for Saudi Arabia as intermediates and then to humans.

• COVID-19 has been supposed to start from the Wuhan local market (Hubei province - China) in December 2019. It may also come from bats which have transmitted the virus to the pangolin sold in the Wuhan market and then the pangolin has transmitted to human. However, the origin of the transmission is not quite clear and there are some hypotheses that the virus may have been invented inside the Wuhan research laboratory and has been propagated inadvetently. The transmission mode from humans to humans is through saliva droplets, sneezes, coughing and contacts.

• The virus has subsequently propagated to the rest of China and contaminated up to now (15/04/2020) 82 295 persons. Today, allmost all countries of the world are now affected leading to 1 997 321 infected persons with 500 819 cured and 127 601 deceased. The 5 countries with most affected persons relative to the total number of cases and to the population density are shown in Tables 1 and 2, where one can notice that the relative number per million of infected persons is very high in small area countries.

**Table 1:** The top ten countries having the highest numbers of contaminated persons and above 100 000 persons on 08/05/2020. The data come from [5].

| Rank | Country | Absolute # | # per million |
| --- | --- | --- | --- |
| 1 | USA | 1 289 235 | 3 912 |
| 2 | Spain | 221 447 | 4 702 |
| 3 | Italy | 215 858 | 3 583 |
| 4 | United Kingdom | 206 715 | 3 112 |
| 5 | Russia | 177 160 | 1 207 |
| 6 | Germany | 169 430 | 2 038 |
| 7 | France | 137 779 | 2 054 |
| 8 | Brazil | 135 773 | 642 |
| 9 | Turkish | 133 721 | 1 608 |
| 10 | Iran | 103 135 | 1 238 |

**Table 2:** The top ten countries having the relative highest numbers of contaminated persons per million of population on 08/05/2020. The data come from [5].

| Rank | Country | # per million | Absolute # |
| --- | --- | --- | --- |
| 1 | San Marino | 18 526 | 622 |
| 2 | Andorra | 9 698 | 752 |
| 3 | Qatar | 6 876 | 18 890 |
| 4 | Luxembourg | 6 286 | 3 859 |
| 5 | Island | 4 944 | 1 801 |
| 5 | Spain | 4 702 | 221 447 |
| 6 | Ireland | 4 548 | 22 385 |
| 7 | Belgium | 4 462 | 51 420 |
| 8 | Gibraltar | 4 273 | 144 |
| 9 | Guernsey | 4 013 | 252 |
| 10 | Singapore | 3 671 | 20 939 |

• COVID-19 then appears as the most devastating pandemic of the 21th century^1^. To face this unexpected drama where there is no known medicine^2^ and vaccine^3^ to fight the virus, only urgent preventive measures have been taken by different countries.

• As the virus is quite heavy, it cannot travel more than 1 meter^4^. Its lifetime is relatively short on skin (few minutes) but long in air and copper (3-4) hours, clothes (12 hours), cartons (24 hours), woods, plastics and metals (3-9 days)^5^. So different preventive barriers have to be used (masks, soaps, hydroalcoholic gels,..) to stop the virus.

• However, in addition to these barrier precautions, drastic measures such as confinement have been taken by governments of different countries which uneluctably affect the social organizations and economic situations. Confinement may lead to a new form of society and economy, to a new way of leaving in the near future.

• Different analysis of the propagation of COVID-19 appearing in China, Europe, UK, USA and Russia have been done in different works (see e.g. various new articles in arXiv [9]) where most of them are based on Susceptible-Infected (SI) or Susceptible-Infected Recovered (SRI) models (see e.g. [10–21]) while some is a simple Gaussian fit of the data [22]. To our knowledge nothing has been yet done in African and some other developing countries. The reasons could be that, in these developing countries, the pandemic is still at the beginning of its effect and the data are not yet sufficient to make a rigourous statistical study (Gaussian law of large numbers of events) which is also complemented by the eventual non-reliability of the collected data due to the few numbers of detection tests or perhaps for some political reasons.

## 2. The spread of infected persons

### 2.1. First analysis: data up to 15 days

COVID-19 enters in Madagascar on 21th March through the Antananarivo-Ivato airport via passengers from Europe and from China. The official measure for closing the airport was too late despite the requests of many individuals or through the social networks.

• We collect, in Table 3, data from different national newspapers [2] and TV [3] issued from the national representant [4] of the World Health Organization (WHO) / Organisation Mondiale de la Santé (OMS).

**Table 3:**
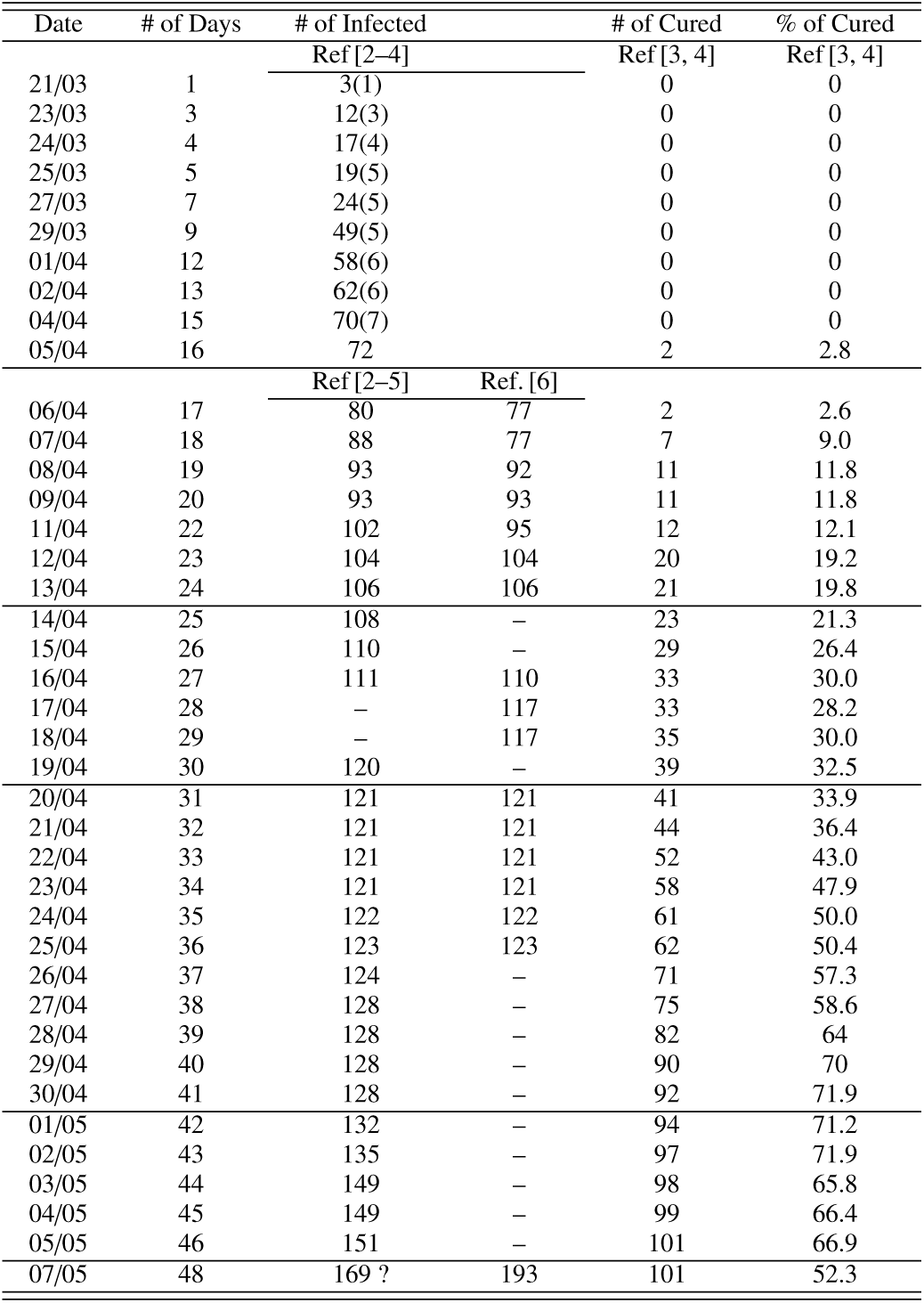
Numbers of Contaminated Persons in Madagascar at different dates. The data from 21/03/ to 15/04/ have been communicated by the national press [2] and TV [3] issued from the WHO / OMS agency in Madagascar [4] and from Google [5]. Data compiled by WHO / OMS are from [6]. The added quoted errors are an estimate of about (10-20)% systematics which can be an underestimate. The 2 last columns are the number and percent of cured persons.

• We use the data of the first 15 days to predict the ones from 15 to 20 days. Noting that some data do not agree each others and that the detection tests are only done for few samples of the population, we have introduced some estimated errors in order to quantify this deficit.

• The analysis is shown in Fig. 1 where the input data are the green circles. Our prediction (filled region) comes from a standard least-*χ*^2^ fit of the data using a Mathematica Program elaborated by [7]. The central value of the data is fitted by the lowest curve while the central value ⊕ the estimated errors by the upper curve. The fit is extrapolated until 20 days.

**Figure 1:**
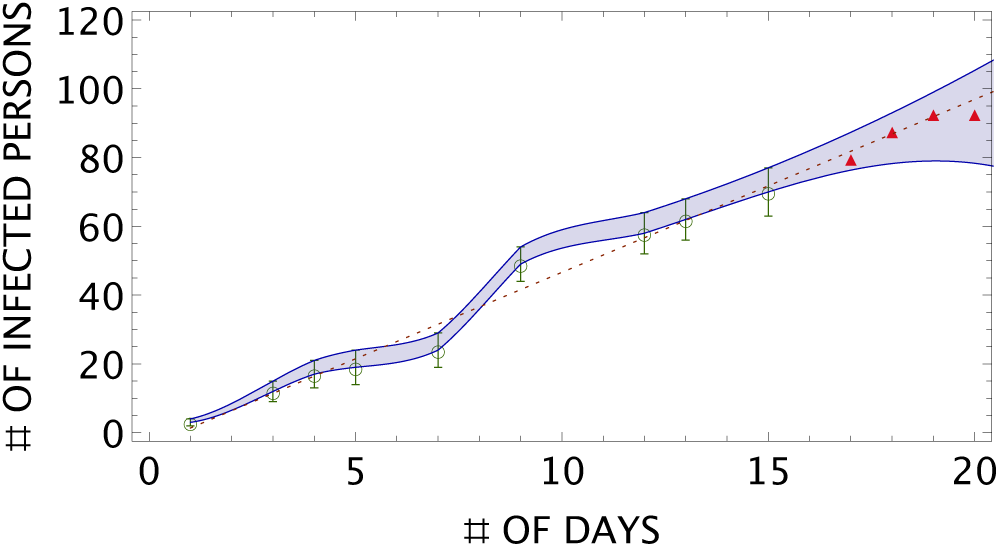
Number of contaminated persons versus the number of days: green circles are the input data, filled region are the predictions limited by the lower curve (central value of the data) and by the upper curve (data with added positive estimated errors). Dashed red line is the linear fit. Triangles are data received after the fitting procedure.

• The new data points in red are inside our prediction region which we consider as an encouraging positive test of the approach based on a standard least-*χ*^2^ fit of the data.

• We also show (dotted curve) the expectation from a simple linear parametrization of the data as given by:

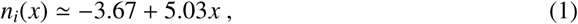

where *n_i_(x)* is the number of infected persons as function of the number *x* of days. The data show a linear increase for about 5 persons per day.

### 2.2. Second analysis: data up to 30 days

• Encouraged by this positive test, we improve the analysis by adding the new data (see Fig. 2) collected from the 16th to 30th days.

• The, *χ*^2^-fit is shown by the continuous green line (green circle data from [2, 5] and dot-dashed red curve (red triangle data from [6]).

**Figure 2:**
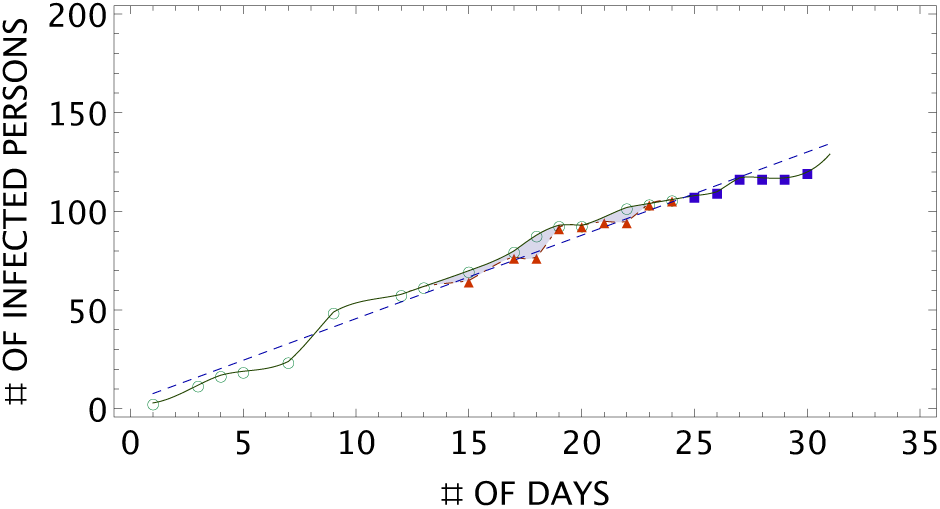
Number of infected persons versus the number of days: green circles are the input data from [2, 5]. The red triangles are data from [6] from the 17th day. Blue rectangles are new data [2, 5, 6] from 25th to 30th days. The continuous oliva curve is the fit from 1 to 30th days of[2, 5] data while the red dot-dashed one is the fit of [6] data which follows the red triangles data points. The dashed blue line is the linear fit given in Eq. 2.

• Doing a linear fit of the data collected during 24 days, we obtain:

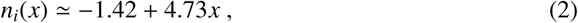

while adding the data until 30 days (blue rectangles in Fig. 2), we obtain:

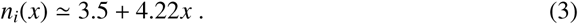

The slopes of the linear curves obtained in Eqs. 2 and 3 ((blue dotted line) are consistent with the one in Eq. 1 shown in Fig. 2. This linear fit can be improved by using a quadratic fit of the data. In this way, one obtains:

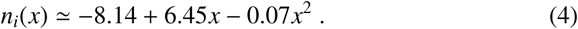

The fits indicate an average of about (4-5) persons infected per day.

### 2.3. Comments on these results

• The previous analyses have been done using data from [2, 5, 6]. We are aware that these data may be inaccurate as there are some periods (week-end) where the data remain stable which may due to the break of the infection tests service. These data may also underestimate the real number of infected persons due to the insufficient number of detection tests for the whole country which contains about 80% of rural population (see e.g[24]). The recent news [2] indicates that about 2200 tests have been performed. The detection tests should be extended for improving the quality and reability of the analysis.

• However, despite these warnings, the reported data can be considered as lower bounds of the real case and the results from the previous study may already be useful. The results of the analysis can be an important guide to the government for taking the right decision at the right time in order to control this tremendous pandemic.

• Our analysis concerns only the officially declared contaminated cases and does not subtract the numbers of cured and deceased persons.

• However with the last collected number (19/04/2020) for about 2 200 tested persons [2] which are relatively small compared to the 27 million of population, one has [5, 6] 120 declared cases, 35 cured persons and 0 deceased. One can also notice that the number of cured persons increases linearly for about 2 persons per day.

### 2.4. Comparison with some other approaches

• Our results from the,*χ*^2^-fit (continuous oliva line), from a linear (dashed blue line) and a quadratic (dotdashed green curve) parametrizations of the data are shown in Fig. 3.

**Figure 3:**
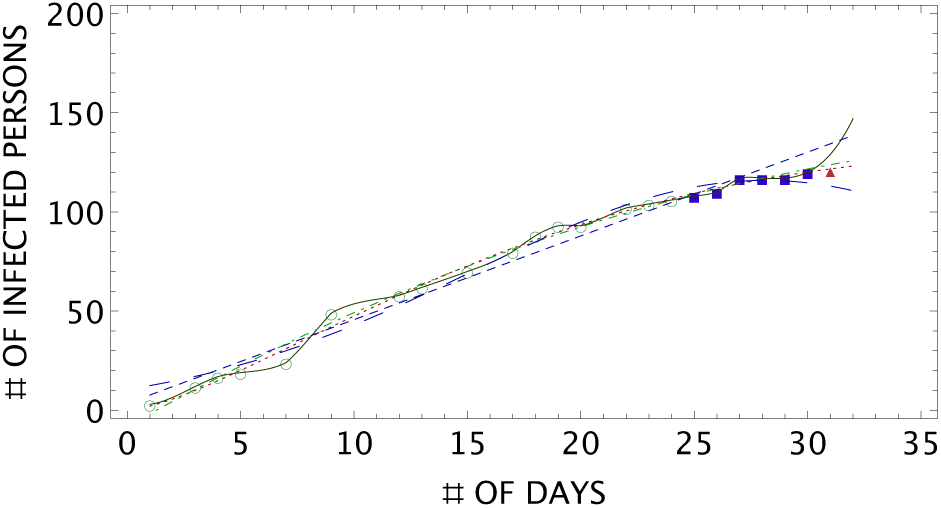
Data are the same as in Fig 2. New data (red triangle) is not included in the fit. The continuous oliva curve is the fit from 1 to 25th days of [2, 5] data and 25th to 30th days of data from [2, 5, 6]. The dashed blue line is the linear fit in Eq. 3 while the dot-dashed green curve is the quadratic fit given in Eq. 4. The dotdashed red curve is the fit from the SI-like model used in [10, 11] while the large dashed green curve is the one from a guassian-like fit used in [22].

• We shall compare your previous results with some from different approaches in Refs. [10, 11, 14, 15, 17, 18, 20–22], though we are aware that these models are more accurate for a large number of events.

• Among these different models and for an illustration, we choose the one in [10, 11] using a variant of the (SI) model with mixed power and exponential behaviours [23]:

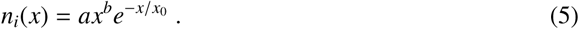

The data are fitted by the parameters: *a =* 2.2, *b =* 1.5 and *x*_0_ ≈ 28 days which we show in Fig. 3 (small dashed red curve). One can notice a good agreement between the quadratic polynomial fit and the one from the SI-like model of [10, 11].

• We also fit the data using a Gaussian-like function as in [22]:

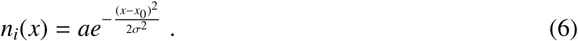

The result is shown in Fig. 2 (large dashed green curve) for a gaussian centered at *x*_0_ ≃ 28 days and a=115.9, *σ* = 12.8.

• One can notice that all different models indicate a slow increase of the number of infected persons which can be checked by more forthcoming data. Our Model with polynomial behaviour leads to a good fit of the data. However, the decrease shown by the Gaussian model may be unrealistic.

### 2.5. Extension of the Analysis until 46 days

• During the submission of the paper, new data based on 3700 detection tests appear (see Table 3), where one can observe an almost stable number of infected persons from 30 to 42 days which requires a reparametrization of the different models. Therefore, we extend the analysis until 46 days. The results of the analysis are summarized in Fig. 4.

**Figure 4:**
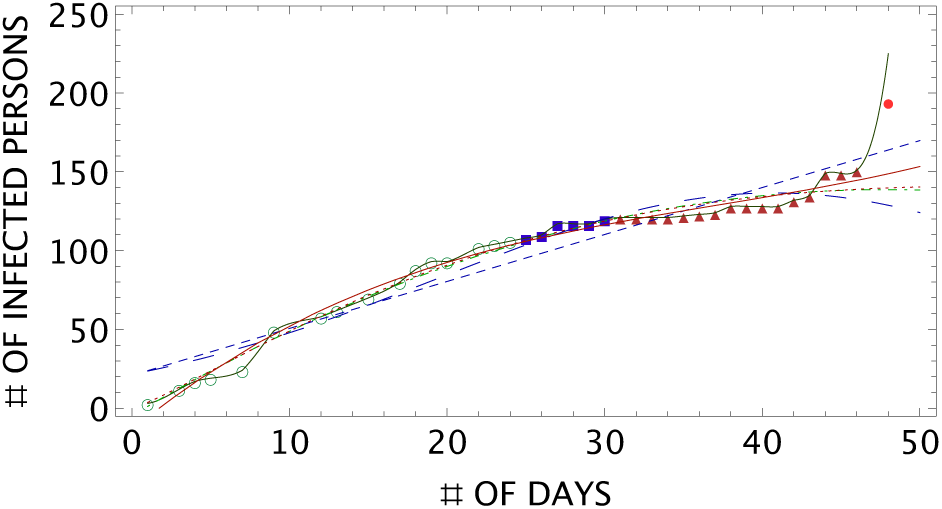
Previous data are the same as in Fig 3. New data (dark red triangles) are added in the analysis.The dashed blue line is the linear fit in Eq. 3. The dot-dashed green curve is the quadratic fit given in Eq. 4. The full red curve is the cubic fit. The dotdashed red curve is the fit from the SI-like model used in [10, 11] while the long dashed blue curve is the one from a guassian-like fit used in [22]. The continuous oliva curve is the one from of least *χ*^2^-fit. The red point is the new data on 7th may not included in the fit.

• One can see from there that the mixed power-exponential model used in [10, 11] (dot red curve with *a* = 3.74, *b* = 1.22, *x*_0_ = 44.08) and the quadratic polynomial fit (dashed blue curve with *n_i_(x)* = −4.81 + 6.00*x −* 0.06*x*^2^) give almost the same predictions, while the gaussian-like model used in [22] (large dashed grey curve with *a* = 136.58, *x*_0_ = 40.73, *σ* = 21.19) fails to describe the most recent three data. The linear fit provides a good smearing of the evolution number:

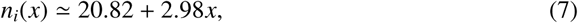

indicating an average of about 3 infected persons per day. The best fit is obtained from the cubic polynomial:

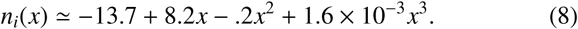

This cubic polynomial model signals a future sharper growth while the other models predict some future stabilities or even a decrease of the number. However, these features from these other models look unrealistic as the virus incubation period of about one month is just finished while the winter season is approaching which could be a favoured period for the virus.

**Figure 5:**
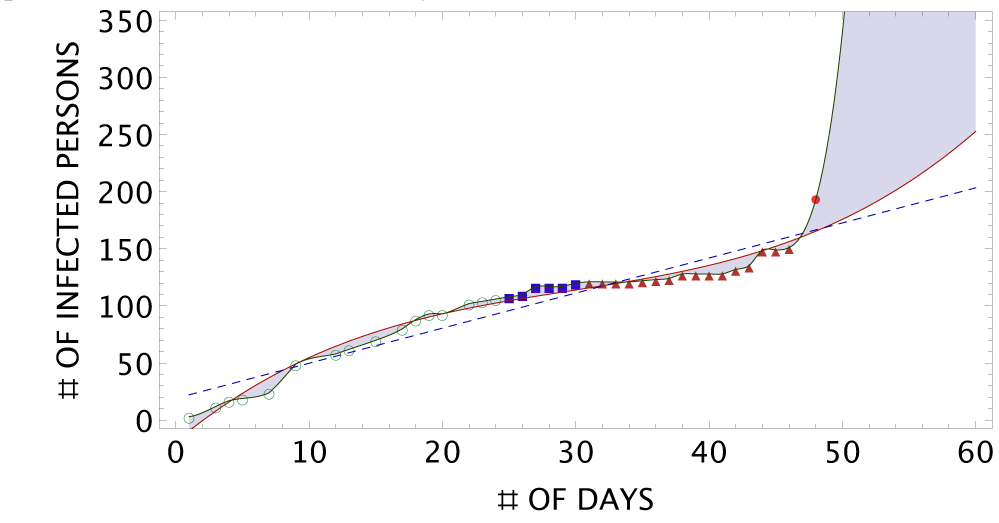
Data are the same as in Fig 4. New fit including the new data 193 (red point) on 7 may from [6]. The oliva curve is the *χ*^2^-fit. The continuous red curve is the cubic polynomial. The dashed line is the linear fit.

### 2.6. Data of the 48th day as a signal of a new phase

• We show in Fig. 4 the new data of the 48 day based on 3968 detection tests. It indicates a sudden jump compared to the previous data such that the different models successful to describe the data for the first 46 days fail here.

• However, the new data (red point) agree with the least *χ*^2^-fit method used to fit the former data. This feature may signal a new phase for the spread of the infection. It coincides with the early partial deconfinement, the end of incubation period from contacts and the beginnng of the winter time where the UV destruction on the virus may be less efficient.

• From the previous analysis, we expect that the most conservative predictions for the next days are inside the region (grey colour) limited by the cubic polynomial (red) and least-*χ*^2^-fit (oliva) continuous curves. Note that the new data 193 from the WHO site [6] has been revised downards to 169 ones from the OMS national representant [4]. However, this change does not affect the qualitative behaviours of the two curves though decreasing its absolute value by about 20.

## 3. Evolution number of cured persons

### 3.1. Analysis until 38 days

• We complete the paper by analyzing the evolution of the cured persons until 38 days. Fig. 6 shows the total number of cured persons per number of days. One can notice that the three parametrizations (linear and quadratic polynomials and the mixed power-exponential model) provide a good description of the data with the forms:

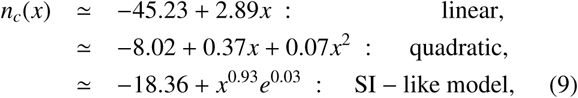

where we have added a constant term for the SI-like model in order to take into account the fact that during the first 15 days there were no cured persons.

**Figure 6:**
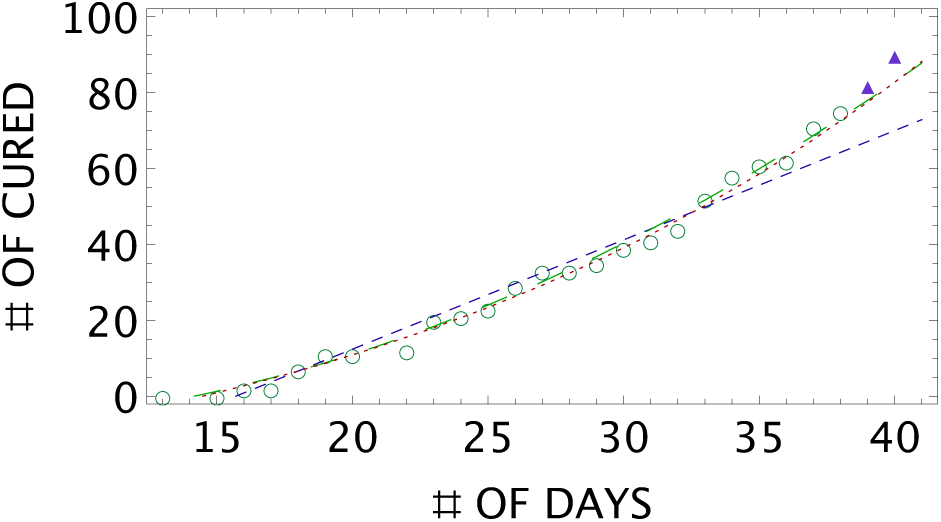
Comparison of different model predictions using the data in Table 3. The dashed blue line is a linear fit. The long dashed green curve is a quadratic fit. The dot red curve is the prediction of the modified SI-like model in [10, 11]. Green circles are data until 38 days. Blue triangles are new data for 39 and 40 days not included in the fitting procedure.

• Fig. 7 shows the total percentage number of cured persons per number of days. The behaviour is similar to the one in Fig. 6 and is parametrized as:

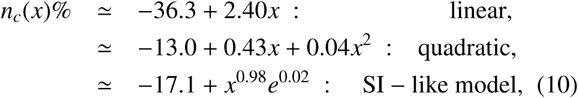

**Figure 7:**
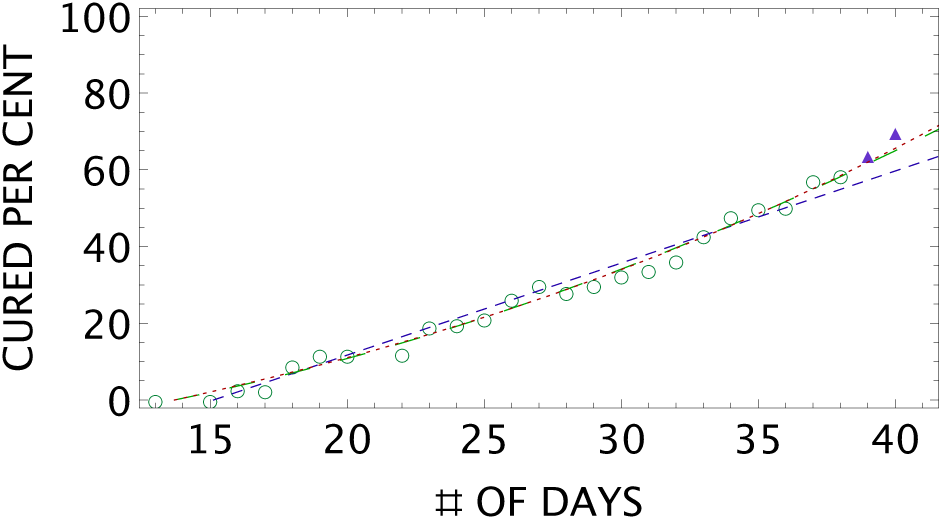
The same as in Fig. 7 but for the per cent number of cured persons. The data fromTable 3 until 38 days are the green circles. Blue rectangles are new data for 39 and 40 days not included in the fitting procedure.

• From this analysis, one can deduce from the linear fit an average number of about 3 cured persons which correspond to about 2.4% of the total number of declared infected persons.

### 3.2. Analysis until 46 days

• We pursue our study by analyzing the evolution of the cured persons until 46 days. The data are shown in Table 3. We compare the predictions of different models in Figs. 8 and 9.

**Figure 8:**
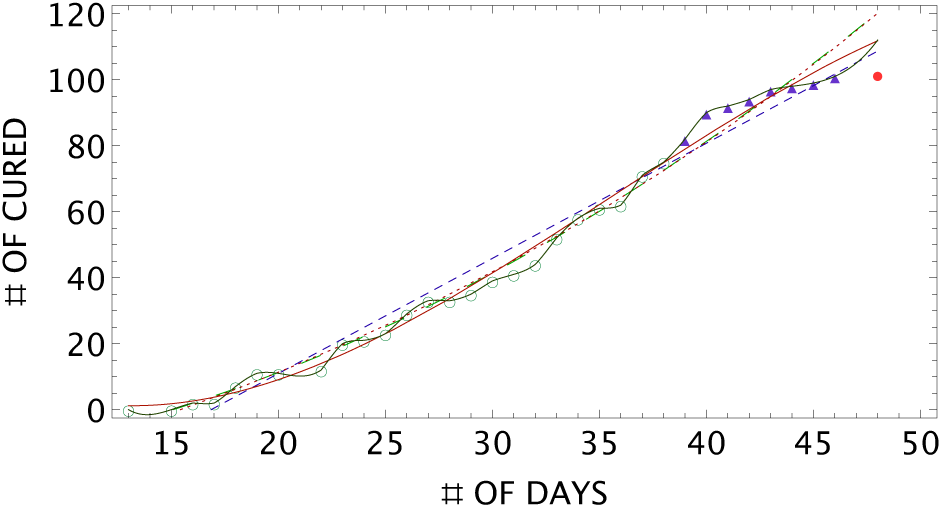
Comparison of different model predictions using the data in Table 3. The dashed blue line is a linear fit. The long dashed green curve is a quadratic fit in Eq. 4. The dot red curve is the prediction of the SI-like model in [10, 11]. The continuous red curve is the cubic polynomial fit. The continuous oliva curve is the result from a least *χ*^2^-fit. Green circles are data until 38 days. Blue triangles are new data until 46 days. Red points are data from 48 days not included in the fit.

**Figure 9:**
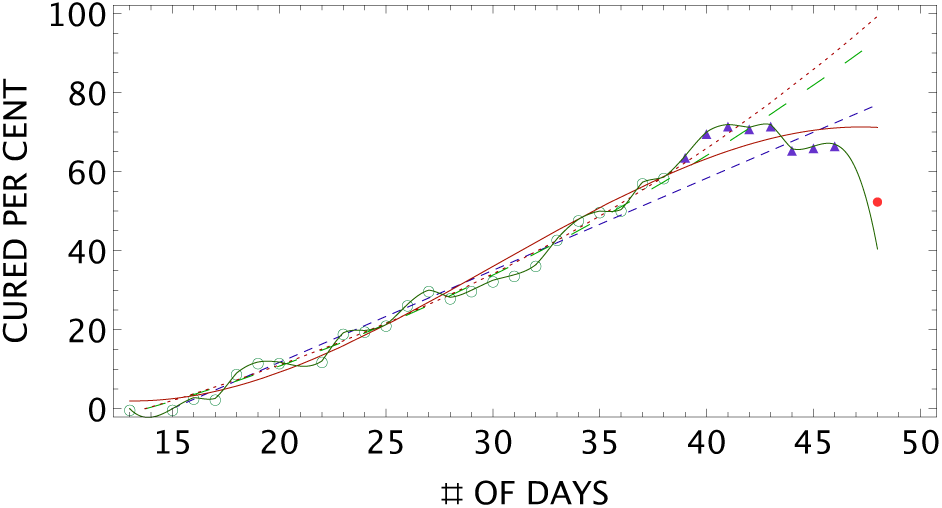
The same as in Fig. 8 but for the per cent number of cured persons. The data come from Table 3. Green circles are data until 38 days. Blue triangles are new data until 46 days. Red points are data from 48 days not included in the fit.

• One can see from these two figures that the best fit including the last three data is obtained from the linear and cubic polynomials. For the total number of cured persons, they read:

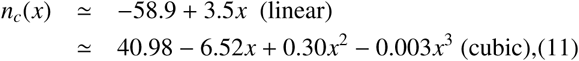

which indicates an average of about 3.5 cured persons per day.

• For the percentage number, the result is:

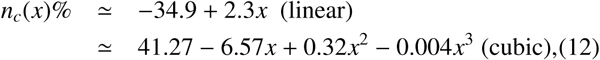

which is about 2.3% of the total number of infected persons.

• The quadratic fit (long dashed green curve) and the model used in Ref. [10, 11] give almost the same predictions but fail to reproduce the last three data points. Their corresponding parametrizations for the total number of cured persons are:

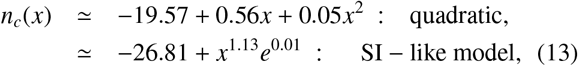

and for the percentage:

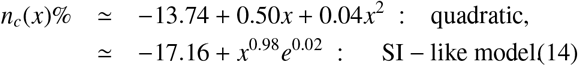

• One can observe the relative good performance of the medical care (2.9-3.5) cured persons per day compared to (4-5) infected persons per day and that no deceased persons have been officially declared since the beginning of the pandemic.

### 3.3. Analysis beyond 48 days

• One can see from Fig. 8 that the new data from the 48th day on the total number of cured persons are better predicted by the linear and cubic polynomials as well as by the least,*χ*^2^-fit used to analyze the data below 46 days.

• However, the percentage number of cured persons is only predicted by the least, *χ*^2^-fit as can be seen in Fig. 9. It indicates a relative decrease of the care performance which is due to the sudden growth of the number of infected persons.

## 4. COVID-19 and Confinement for Madagascar

### 4.1. Political decisions

As already mentioned in the introduction, in order to limit the spread of this dangerous virus which propagates via social contacts (coughing, saliva droplets,..) and waiting for a new efficient medical drugs, different countries have decided to confine the population and have asked the persons to strictly respect some barriers rules. About 3 billions of persons i.e half of the humanity are now / have been confined.

### 4.2. Concretization

However, the concretization of confinement in developing countries is far to be achieved due to poverty and to the lack of education of the majority of the population:

• Due to poverty and to the bad organization of the society, most of persons have to work for finding what to eat day by day due to the informal forms of the trade, business and economy.

• In addition, the accompanying help measures taken by the government are insufficient while the managements of some international funds and donations are not transparent.

• Middle class persons also suffers as they are not rich enough to be autonomous and not too poor to receive any help from the state.

• Due to the lack of education, most of the people are irresponsable and are not aware about the dangerous effect of the virus. Then, they do not see the importance and do not feel obliged to respect the barriers measures and the confinement.

## 5. Worldwide Impacts of COVID-19 and Confinement

As a consequence, these unprecedented pandemic and confinement security measures have large impacts for the:

### 5.1. Social organisation

• Most of us learn about teleworks, indoor at outdoor houseworks.

• We re-discover the importance of a family and of the tradition

• We re-discover ourseleves from our concentration and meditation.

• We see the usefulness of solidarity.

• We see the values of health personals, researchers, teachers, educators. firefighters,…and in general the human values.

• Urban exodus in developing countries such as Madagascar are observed.

• However, the pandemic might enhance the social class inequalities like the increase of the difference between poor and rich peoples, the re-disappearance of middle class, mentioned earlier for Madagascar, which is the lungs of developments.

### 5.2. Environments

• Nature takes back its rights: returns of animals near cities and cetaceans near the coasts, returns of insects, birds,…).

• Air pollution is decreasing due to a minimal road traffic. Megacities (Beijing, New-Delhi, New-York, Parish,…) recover an improved atmospheric air.

• Cities become less noisy.

…

### 5.3. Economy

• Globalization is suffering.

• Reduced size and local markets are developing.

• Small producers are reorganizing.

• On-line sales and drive markets are developing.

• Delocalisations of manufacturers and factories are questioned.

• Each country is looking for an independent and self-sufficient economy which may lead to a protectionism.

### 5.4. Politics

• COVID-19 has pressed the leaders of each country to revisit the orientation of their politics to the most useful ones for the population namely:

– Health,
– Research and Education,
– Foods and different ways of Consuming,
– Protections of the Environments.

• COVID-19 has stimulated the search of an autonomous country which, if not done carefully, may lead to an ultranationalism and to a withdrawl into oneself (closing of boarders,…)

## 6. Summary and Conclusions

• We have studied the first month spread of COVID-19 in Madagascar using standard least *χ*^2^ fit approach. We found that, for this first month, the spread of the virus per day increases almost linearly/quadratically for about (4-5) persons per day. A comparison of our results with the ones from some other SI-like models is shown in Fig. 3.

• The inclusion of the recent data from 28 to 46 days indicates new features which are essentially due to the plateau from 30 to 40 days (see Fig. 4) and requires a new parametrization of the data. Indeed, these additional data favour a cubic polynomial fit instead of the quadratic one and of the models used in [10, 11,22].

• These data do not indicates any sign of an exponential growth of the number of infected persons though the cubic polynomial signals a stronger increase in the near future.

• Indeed, the recent data collected beyond the 48th days show a sudden jump which are not explained by the previous models except the least *χ*^2^ fit as shown in Fig. 4. This sudden jump signals a new phase on the spread of the virus which may coincides with the end of the incubation period (contact case), the arrival of the winter period which the virus may like and with the beginning of the partial deconfinement. We show in Fig. 5 the new fit including this new data. The prediction for the next days are given by the grey region limited by the, *χ*^2^-fit (oliva curve) and the cubic polynomial (red curve).

• We note that the number of infected persons are relatively low, which can be due to the reduced number of detection tests. Another factor could be that the intense UV in the country during this first month may have partially neutralized the virus effects.

• We have also analyzed the number of cured persons where we note that its behaviour is analogue to the case of infected persons. The linear/quadratic polynomial fit and the SI-like model with mixed power-exponential behaviours fit quite well the data before the 40th days but, with the inclusion of the new data from 38 to 46 days, the whole data are better fitted by a cubic polynomial like in the case of infected persons.

• One has also found that the percentage number of person cured corresponding to the new data beyond 48 days are not reproduced by the different models discussed above but only by the least ^-approach which signals a relative decrease of the percentage care as a consequence of the sudden increase number of infected persons.

• It is remarkable to note that the total number of about (2.9-3.5) officially cured persons per day relative to the number (4-5) of infected persons is a good performance despite the poor equipment of the hospitals. Then, one wonders if these care techniques are based on modern chemical drugs or complemented by some endemic medicinal plants ? More explanation, on the technical cares used to get these succesful results, is greatly appreciated. One should also note that our study has been done before the official distribution/sale of the CVO based artemisia tea claimed by the governement to cure/prevent the COVID-19 pandemic but this claim is under debate.

• We have also noticed the relatively low number of tested persons which is around 3 700 persons (date 04/04/2020) compared to the total number 27 millions of population where 1.6 millions are in the capital Antananarivo. A large effort for increasing this number of detection tests, its origin and the kind of samples is required to have a more realistic basis for a more robust scientific analysis.

• More transparency on the uses of the international donations for the hosptial equipments, materials for detection tests, barrier material (masks, hydroalcoholic gel,…), the help for the poor peoples is continuously requested.

• We have shortly reviewed the general Worldwide impacts of COVID-19 which has demonstrated the weakness of the current mondial global system. This feature might announce a change towards a new model of society and economy and for a new form of political decisions in the near future.

## Data Availability

Data used to get the results are in Table 3 of the paper where references to the sources can be found.

## Acknowledgements

It is a pleasure to thank S. Maltezos, R. Narison and R. Ziff for reading the manuscript and for some communications.

This research has not received any dedicated funds and sponsoring. It is issued from a personal initiative without any influence from private and public organizations.

## Footnotes

1 For a more complete but simple review on the cornavirus, see e.g. [8].

2 Drugs based on chloroquine proposed by Pr. Didier Raoult - Marseille (expert in tropical diseases), which were successful against paludism are promising though under debates. Some other pists are the uses of some endemic medicinal plants such as in Madagascar where some of them contains natural chloroquine.

3 There is some proposal from the Bill Gates foundation but doing the first tests in Africa are criticized.

4 However, new study (French TV source) recommends a distance above 2m.

5 The true lifetime of the virus on inert objects is still under study and needs to be confirmed.

